# Professional inclusivity: Creating faculty development learning environments to facilitate learning across the health professions

**DOI:** 10.1101/2025.05.02.25326878

**Authors:** Maura N. Polansky, Jascha de Nooijer, Goetz Fabry

## Abstract

**Background:** Learning environments involving multiple health professions may provide opportunities for interprofessional learning (IPL). However, not all multi-professional settings provide the necessary conditions to support IPL. Using the concept of professional inclusivity, the purpose of this study was to explore conditions that support professionally inclusive learning environments in faculty development, using the example of Master in Health Professions Education (MHPE) programs.

**Methods:** Semi-structured interviews with 14 students and faculty from four MHPE programs were conducted. Member checking of key themes was performed to enhance validity.

**Results:** Organizing principles for supporting professionally inclusive learning environments were identified: (1) being intentional, (2) leveling the playing field, and (3) focusing on commonalities. Intentionality refers to the essential role of faculty in considering how each aspect of the program can impact inclusivity. Leveling the playing field refers to establishing a culture where students (and often faculty) are seen as equal regardless of profession. Commonalities relates to the common background of students, in both clinical and teaching experiences, and their shared educational needs.

**Conclusions:** Organizing principles for fostering professional inclusivity in faculty development learning environments were identified. Specific recommendations for the application within MHPE programs are provided. Professional inclusivity may be of significant valuable as a conceptual framework for further research regarding IPL.

## Introduction

Interprofessional practice is deemed as essential to advancing clinical and academic outcomes.^1-3^ For more than a decade, programs across the health professions education continuum have been called upon to prepare future and current healthcare professionals to work collaboratively with other professions. Much of this work has focused on interprofessional education (IPE). IPE is a pedagogical approach to developing competencies for collaboration involving learners from two or more professions who learn with, from, and about each other.^4^ While IPE is likely a key approach to the development of competencies for interprofessional practice, it is recognized that interprofessional learning (IPL) may result from a variety of educational approaches, including experiences that do not meet the strict definition of IPE.^5^

Learning environments that include participants from multiple health professions may provide important opportunities for interprofessional learning (IPL). However, not all multi-professional educational environments facilitate learning across the health professions.^1^ Understanding the conditions that create a conducive environment is likely key to establishing effective IPL experiences.

Recent scholars have highlighted the important role of educators and leaders in advancing interprofessional learning and practice.^6^ Since attention to interprofessional competency development is rather new, it is likely that those currently in or aspiring to educational leadership positions do not themselves sufficiently possess competencies for effective interprofessional clinical and academic practice. Therefore, professional development programs for educators and leaders in health professions education (HPE) may play a critical role in ultimately preparing the next generation of healthcare professionals to be effective members of interprofessional teams.^7-8^

Continuing professional development programs awarding a master degree for educators and leaders in the health professions have existed for more than 60 years.^9^ These offerings have recently expanded, with an increasing number of degree programs in HPE, most notably those at the master’s level (MHPE).^9-12^ Many of these programs enroll students from various health professions, resulting in multi-professional learning environments and thus, may offer an important opportunity to develop educators, leaders, and scholars who are uniquely qualified to support IPL among students and trainees within their own professional contexts.

Several scholars have followed trends in the expansion of MHPE programs, although the interprofessional nature of these programs has not been a focus of investigation.^9-10,13^ In fact, a comprehensive literature search identified only one study, evaluating a single MHPE program, which aimed to determine how well the program achieved its learning outcomes, including developing interprofessional competencies.^14^ While study participants reported an “overwhelming” positive response to the program meeting its educational aims, the investigators did not attempt to understand what practices or conditions were perceived as beneficial to this IPL. Therefore, no empirical evidence is available to understand the conditions within a multi-professional MHPE program that supports IPL.The literature on IPL (primarily regarding interprofessional clinical practice) highlights the importance of breaking down barriers, reducing myths and misconceptions of other professions, and creating an “open and trusting environment” for learning.^15^ But how educators can create such an environment is under-studied and inadequately theorized and is the focus of this study.

For our work, we identified the construct of “inclusivity” as a suitable conceptual framework to study the interprofessional learning environment of MHPE programs. The term “professional inclusivity” has been previously used (although not formally defined) in the HPE literature as it relates to how students are socialized within their respective professions.^16^ However, socialization within one’s own profession does not necessarily equate to professional socialization within an interprofessional context.^5-6, 15^

While inclusivity as it relates to professional identity is not currently reflected in the IPE literature, it has been explored in other educational contexts, such as the inclusion of racialized learners and others from marginalized backgrounds. Scholars have suggested that being integrated into a learner environment is insufficient for true inclusion.^17^ It has also suggested that an inclusive approach to education must take into account: curriculum design and content; delivery and pedagogy; student assessment and feedback; and institutional commitment to and management of inclusive learning and teaching.^18^ Furthermore, leaders are seen as essential in establishing inclusive environments. Therefore, faculty, as leaders in the classroom, must demonstrate leader inclusiveness which has been described as “words and deeds by a leader or leaders that indicate an invitation and appreciation for others’ contributions”.^19^ Furthermore, to ensure educational excellence by faculty, their practices must support a sense of belonging; view differences as opportunities for learning; and ensuring that educational cultures, policies and practices reflect a valuing of everyone equally.^17^

For the purpose of this study, we propose the following definition of professional inclusivity as *a learning environment where participants from differing health professions are respected, valued, and supported to facilitate interprofessional learning*. The aim of this study is to explore the conditions that support a professional inclusive learning environment in MHPE programs.

## Materials and Methods

### Participants

This study involved interviews with faculty and students/alumni from U.S. MHPE programs. Individuals were recruited from four programs, purposefully selected based on information on their website indicating they were actively enrolling students from two or more professions. Programs represented were from different geographical regions of the U.S., including both public and private institutions. Curricula were primarily delivered online/remotely, with limited in-person activities. No program represented in the study was delivered entirely in-person.

Participants were recruited through published contact information or individuals familiar to the research team. Program faculty were contacted directly via their institutional email. Potential participants who were current or former students were contacted via email through the faculty participants. Interested individuals who expressed interest in participating, recieved an informed consent document via email.

### Research team

The research team included three investigators who have direct experience as faculty and former students within MHPE programs. They also have professional expertise working in interprofessional settings.

### Interview guide

Interview guides (provided as a supplement) were developed for students/alumni and faculty and designed to explore participants’ experiences in and perspectives of the MHPE program. Topics included relationship building and behaviors influencing a sense of belonging, educational activities of the program and its curriculum, drawing on concepts that align with the before mentioned literature on inclusivity.

Information including professional background, affiliated program, and current career status was collected at the beginning of each interview to provide context for the analysis.

### Data collection

Participants were interviewed by the primary investigator (MNP). Interviews were conducted and recorded via a web-based platform (i.e., Zoom). The Zoom transcription feature was used to create an initial transcript. A research assistant reviewed the recordings for accuracy of the Zoom transcript, resulting in verbatim transcripts. Second, the de-identified versions were prepared for analysis.

### Data Analysis

Two investigators (MNP & GF) reviewed all transcribed interview data. Line-by-line coding was initially conducted and key findings were identified through an iterative process. Investigators met regularly to discuss key themes identified in the data. They participated in reflexivity, including through reflecting on their own prior experiences as students and instructors in similar learning environments.^20^ After key findings were identified and guiding principles developed, they were finalized by the full research team. Theoretical sufficiency was determined through these team discussions.^20^

As part of member checking^20-21^, all participants were contacted via email with key findings from the analysis and asked to comment on the primary findings to determine if they were felt to capture their perspectives accurately. Seven participants responded providing agreement with study findings, with a single point of clarification provided by one participant.

### Study ethics

The protocol for this study was deemed exempt from institutional review board review by the University of Illinois, Chicago.

Given the small number of MHPE programs in the U.S., only limited participant data in aggregate is reported to ensure anonymity of participants.

## Results

Fourteen participants (8 faculty and 6 students/alumni) were enrolled in the study. Participants represented different professional backgrounds, including medicine (physician and physician assistant/associate), nursing, psychology, medical imaging, basic sciences, and educational administration.

### Organizing principles for professional inclusivity

The primary findings from our analysis related to organizing principles that appeared to be essential to developing a professionally inclusive learning involvement. These three principles are (1) being intentional; (2) leveling the playing field; and 3) focusing on commonalities.

Participants emphasized the importance of the program and faculty *being intentional* in creating a professionally inclusive learning environment where all students belonged. Program faculty valued the interprofessional nature of the program and were purposeful in creating a sense of belonging among students. This intentionality related to a variety of aspects of the program including student admissions, faculty recruitment, curricular planning, and providing opportunities for engagement.

#### Leveling the playing

field between students was deemed essential to creating a culture that valued students across different health professions. This principle applied to creating an educational culture where students (and often faculty) were seen as equal regardless of profession or level of prior education (e.g. master vs. doctorate degrees). Another aspect of this principle referred to leveling hierarchies that often are present in clinical and academic environments.

#### Focusing on commonalities

was viewed as essential to creating a sense of belonging. Participants recognized that although students enrolled in the program came from different professional backgrounds, commonalities also existed, e.g., most students had both clinical and teaching experience. There was also emphasis on the shared educational needs for all students related to HPE that included the need to develop competencies in teaching, leadership, and educational research.

Each of these principles was apparent throughout numerous aspects of the program, as discussed below.

### Demonstration of principles

#### Program Curriculum

Program curricula focused on topics and skills applicable to all health professions educators. The key topics addressed within the program were in domains of teaching and learning, educational research, and leadership.

> “We learn about the inadequacies of how we teach. And what we wish we were doing better, but we’re learning about those inadequacies because everybody’s got them. And we’re talking about how to try to fix them together.” (faculty)

Assigned readings and educational resources were seen as impacting professional inclusivity. They were identified from multiple health professions sources, as well as outside of healthcare. Programs drew from literature in various profession-specific journals and texts, the interprofessional literature, and disciplines outside of healthcare, such as the social sciences. The use of these educational materials required faculty to be purposeful about drawing from this wide body of literature.

Assignments included discussions, case analyses, group assignments, and individual projects. Discussion sessions and case analyses focusing on topics relevant to all students or with examples from multiple health professions, allowed students to draw on shared teaching and clinical experiences emphasizing their commonalities. While students were expected to discuss topics related to their own individual work, the use of profession-specific jargon was discouraged, and students were expected to provide sufficient professional context so all students could participate equally in discussions. This approach further emphasized commonalities among students and avoided prioritizing one profession over another, leveling the playing field.

> “I think it’s because the assignments are so general and it’s an even playing field for everyone.” (student)

Group assignments also involved applying concepts to topics relevant to all students regardless of profession. Topics were foundational to HPE, and in some cases, would draw on examples outside of healthcare and were therefore agnostic to the health professions. In contrast, individual projects provided a forum for students to apply course content to their own work and context.

While projects, by design, varied based on the students’ professional background and responsibilities, the focus for these projects was seen as highly relevant to all students regardless of profession. Ensuring that group and individual project assignments were relevant to all students was seen as a responsibility of the faculty in planning these assignments.

> “There are a number of, um, activities where we purposefully choose topics that are not in the background of anyone…And it’s interesting, because almost every year people start saying ‘Why are we doing this in this class?’ And then they come to realize the importance of actually um, you know, getting yourself out of your comfort zone or your area of expertise, and also putting everyone in the same level, whether you are a doctor, or a dentist, or nurse, or whatever.” (faculty)

#### The Hidden Curriculum

Acknowledging the interprofessional nature of the program and encouraging students to learn with and from each other supported the sense of valuing all students regardless of professional background, as part of the hidden curriculum. Being on a first-name basis among faculty and students also supported an inclusive environment, leveling the playing field and flattening hierarchies that often exist in clinical settings.

> “It’s a first name basis program. We’re very big on everybody is equally a member of the community of learning.” (faculty)

Socialization among the student body was also viewed as important. Opportunities for students to make professional and personal connections occurred through various methods, both online and in-person. Student groups were also organized intentionally to provide opportunities for students to work with others from different professions and avoid groups with the majority of students from one profession. “Providing space” in the program, as it related to time in the agenda and physical space during in-person activities for further connection appeared essential.

Early and longitudinal experiences in the program were particularly important in developing a sense of belonging. Using a cohort model or having at least a subset of classes that students take together helped create a sense of belonging. Within classes, being intentional about a balance between longitudinal experiences with the same students and periodically changing group composition gave students a chance to connect with others and build a broader sense of connection across the student community.

#### The Role of Faculty

A critical aspect of each program was the interprofessional nature of the faculty. Each program had faculty from multiple health professions, as well as faculty members from other related disciplines such as social and data sciences. Establishing an interprofessional faculty was deemed essential in creating an educational program and learning environment that was inclusive for all. Recruitment of a professional diverse faculty was seen as a priority, as well as recruiting faculty who valued interprofessional learning and practice.

Faculty having a strong understanding of each students’ profession and its educational model also appeared essential. Co-teaching was an instructional model that provided an opportunity for faculty to learn about other faculty members and their professional backgrounds. Faculty also developed their understanding of other health professions through their engagement with students from different professions through coursework and advising.

#### The Role of the Students

The students themselves were also important in establishing a professionally inclusive environment. It was noted that having a sufficient mix of students from various health professions was important, and therefore, student recruitment was essential. It was also noted that programs should be intentional about promoting their program as relevant to students from various professions. Using recruitment strategies that draw applicants from other professionally diverse programs (such as faculty development programs) and promoting the program at interprofessional conferences was also valuable in these efforts.

Ensuring matriculants have had similar professional experiences was important in leveling the playing field and ensuring that commonalities in their learning needs existed.

> [Classmates of another profession] “are kind of in the same boat we are, learning these skills as well. So, I think it evens the playing field.” (student)

These shared experiences included both clinical responsibilities and efforts in education, with all students seeing themselves as educators. Prior experience of students in IPE matriculants also influenced professional inclusivity. Self-selection was deemed to be an important factor, often influenced by the program reputation or its professional diversity. Similarly, when recruiting students through partners with other faculty development programs that emphasize interprofessionalism, or from educational settings that are typically interprofessional (such as healthcare simulation) matriculating students were seen as prepared for an interprofessional learning environment.

### Interprofessional learning

Although not an explicit aim of this study, some interesting findings regarding IPL were identified in the analysis. While typically not seen an explicit program goal, opportunities for IPL existed within each program and seemed to arise spontaneously. As such, IPL was primarily implicit within the programs and, therefore, not viewed as a formal program goal.

> “[in the beginning] they focused more on theory than necessarily interprofessional education. Interprofessional education as a field, I think kind of emerged or exploded actually after we started our program, which is really interesting.” (faculty)

Some objectives related to interprofessional practice were part of the formal curriculum, making IPL more explicit. The introduction of teamwork and instruction on team science, particularly at the beginning of the program, was also identified some participants. While not typically framed around interprofessional practice, this content was perceived as highly relevant to inter-professionalism. Interprofessionalism was also part of the hidden curriculum such as when faculty from different professions were involved in co-teaching program courses, serving as role models for inter-professional teaching practice.

Some instructional approaches also helped student enhance their understanding of clinical responsibilities of other professions. Since class discussions often required students to apply concepts they were learning to their own context. In doing so, students would reveal information about their profession and its role in healthcare, illustrating important differences in professional responsibilities and competencies where they occur. Similarly, assignments involving students sharing their educational work through presentations to other students or when receiving peer feedback on written projects provided opportunities for other students to learn about their classmates’ professions and at times their educational differences. These opportunistic occurrences provided faculty the chance to explore the professional differences as they arose, seizing on these valuable learning experiences.

> “I think there’s been a couple courses that have made us the teacher and kind of teach our profession a little bit. Which has been kind of, I’ve, I’ve enjoyed, and it’s interesting to learn how other people approach things. “So, for example, one of mine [assignments] was on fetal embryology. So I had like a slideshow about that, and how would I incorporate that in Google classroom. And so then, in that way it-second hand, you learn about folks and how they teach in their professions.” (student)

Finally, the findings show how participants came to value other professions’ clinical and educational expertise. For example, students may demonstrate their extensive clinical expertise while completing a teaching assignment or through peer review activities such as those related to curriculum development.

> “I think that when people get to know about [the student’s profession], I think at first blush it appears to be an easy job, and I use that with air quotes. But the knowledge base that we have, and the skill set that we have, I think folks don’t realize the depth of knowledge of anatomy and pathology that we have to know. And then the … seriousness and responsibility that we have to find pathology and demonstrate it. I think that’s an eye-opener for a lot of folks. (student)

This was also apparent through exposure of students to high-quality instructional materials (e.g. textbooks) from professions other than their own.

> “I don’t think they knew there were nursing journals about education or the way I know they didn’t know there were standards or best practice for simulation. And they’re surprised. And so that, that type of cross pollination happens, and a physician said ‘I’m so surprised that nursing is so developed in this area.’” (faculty)

### Challenges

While a variety of conditions were identified that can support professional inclusivity within MHPE programs, several challenges were also identified relating to student recruitments, physician education “holding more space” (student), and limited in-person experiences.

Although programs aimed to enroll students from various health professions, several barriers to recruitment of a professionally diverse student body were apparent. First was the presumed variability by professions with employers’ support for pursuing advanced degrees. Also some employers and accrediting bodies were noted to prefer faculty to pursue profession-specific educational degrees instead of those that are interprofessional. Ultimately, no program had specific enrollment goals related to the professional background of their student body nor was profession considered to any significant degree in the admissions process due to insufficient numbers of qualitified applicants.

Despite programs efforts to create inclusive learning environments among the health profession, physician education often held more space in the curriculum. Teaching materials and resources were most commonly drawn from medical education and the medical education journals appeared to be the primary source of assigned readings for many classes. Furthermore, topics for course assignments and discussions frequently came from and circled back to medical education.

Finally, in-person experiences were considered superior to online activities in developing connection and a sense of belonging. However, the programs represented were primarily or exclusively online, providing limited opportunities for students to connect with other students and faculty in person. As a means of engagement online, synchronous online activities were sometime used and were perceived as better than asynchronous activities. However, student engagement was further limited when online synchronous and in-person activities were optional.

## Discussion

This study explored the interprofessional nature of MHPE programs and the conditions that support professionally inclusive learning environments within these programs.

Three organizing principles for supporting a professionally inclusive learning environment were identified: (1) being intentional; (2) leveling the playing field; and (3) focusing on commonalities. We discussed how these guiding principles play out through the program as they relate to students, faculty, and the curriculum. Recommendations for other MHPE programs that wish to support professional inclusivity and IPL are provided below.

Professional inclusivity, as defined above, provides an effective lens through which to examine educational practices, and which led to the identification of organizating principles. As such, we think that inclusivity is a valuable conceptual framework in studying IPL environments. While our study specifically explored MHPE programs, these findings may potentially be applied to other multiprofessional learning environments.

Our results show that bringing students and faculty from diverse professions together in a learning environment where students have equal status may help to overcome the traditional hierarchies and power imbalances that typically exist in clinical and academic settings.^22^ Contact theory hypothesizes that participation in diverse groups can reduce prejudice and negative attitudes among its members.^23^ However, mere contact with other professions is insufficient to achieve these results unless equal status among group members exists.^24^ Scholars of IPE further support the notion that flattening hierarchies is key to interprofessional learning within multi-professional groups.^15,25^ Our study provides specific examples of how this can be achieved.

Adhering to these principles helped to create an equal status between students, break down professional silos, and reduce professional misconceptions.^15^ This appears to be key to facilitating the socialization of students within the interprofessional field of HPE.

Consistent with the earlier work by Nembhard & Edmondson^19^ regarding leader inclusiveness, we found that faculty played a critical role (through their “words and deeds”) in developing a professionally inclusive learning environment. Their efforts involved establishing program culture, developing curricula (whether planned, implicit, or hidden), and using pedagogy that reflected a value for and invited participation by students across the healthcare landscape.^17-18^ Additionally, faculty had an important responsibility as role models for collaboration, such as through team teaching, demonstrating how to collaboratively achieve shared goals. The significance of faculty serving as role models for interprofessional collaboration (IPC) has been well-documented.^7,26^ However, instructors may overlook this vital aspect of their role.

Furthermore, role modeling is not an inherent skill (Cimino et al., 2022), and many teachers involved in IPL environments may feel underprepared for their responsibilities.^7,27,28^ Therefore, targeted faculty development, a concept often overlooked in the IPE literature, may be necessary, especially for faculty with limited experience in teaching professionally diverse learners.^29^

### Lessons for practice

The results of this study have led to several recommendations specific to MHPE programs and relevant to the challenges that often exist for these programs.

First, significant representation of different health professions in the student body helps to avoid one group dominating others or having the field of HPE viewed from a single perspective. This can be addressed by practices that support inclusion, such as having promotional materials representing a variety of health professions, using recruitment strategies aimed at diverse professional groups, and seeking commitment from employers for dedicated time and resources for enrollment (such as tuition reimbursement) regardless of profession.

Second, programs should recruit faculty from different professions and disciplines, and with experience teaching in professionally diverse settings; an interest in interprofessional learning; and a commitment to professional inclusion. Faculty development and training should be provided to support those with less experience in interprofessional teaching. When possible, team-teaching and curricular planning with faculty from different professions should be utilized to facilitate the development and implementation of curricula to ensure professional representation across multiple health professions. The curriculum should also draw on the diverse HPE literature, avoiding the risk of one profession dominating the curriculum.

Finally, MHPE programs face unique challenges since most programs are offered online, facilitating the enrollment of health professionals who typically are employed full-time in clinical and academic positions. Programs that have limited in-person contact may face challenges in providing sufficient opportunities for students to establish social connections that support a sense of belonging and community. Therefore, online programs should seek to provide students with opportunities for some in-person courses or activities, when possible.

Alternatively, synchronous online activities that provide time for student engagement could be utilized.

#### Study limitations and future directions

Limitations of this study include limited evidence regarding evaluation data on how the program had impacted competency development in interprofessional practice. The extent of this learning on actual professional practice of program graduates or on their professional identity as members of interprofessional teams is largely unknown (Khalili et al., 2013; Polansky et al., 2022). So, while we suspect that completion of a professionally inclusive MHPE program may impact graduates and their professional practice in meaningful ways, the actual and long-term impact of this unique learning experience requires further study.

Further attention should be placed on the role of faculty and leaders (in both clinical and educational settings) to advance the work of interprofessionalism. While we welcome the efforts that have been made in recent years to develop competencies for early learners in interprofessional practice, more work is needed in faculty development and other type of continuing professional development initiatives (including, but not limited to MHPE programs) to support efforts to transform healthcare into collaborative interprofessional systems.

Further investigation of the concept of “professional inclusivity” is needed. While we found this concept to be quite valuable for this study, it is unknown how it can be applied more broadly in the area of interprofessional education, learning, and practice. Given the robust and emerging body of literature regarding diversity, equity, inclusion and belonging, frameworks and other relevant constructs from this body of work may be of further value in advancing the work in interpofessionalism.

## Conclusion

This study of MHPE programs demonstrated key principles for fostering professional inclusivity among their interprofessional student bodies. The construct of professional inclusivity may be of significant valuable in further research regarding IPL, particularly in continuing professional development programs such as those for faculty in health professions education.

## Data Availability

All data produced in the present study are available upon reasonable request to the authors.

## Acknowledgements

The research team would like to sincerely thank Ms. Ari Buslovich who served as the primary research assistant for this study. We also appreciate the valuable time provided by the study participants.

